# A Convex Optimization Solution for the Effective Reproduction Number *R*_*t*_

**DOI:** 10.1101/2021.02.10.21251542

**Authors:** Joaquín Salas

**Affiliations:** CICATA Querétaro, Instituto Politécnico Nacional

**Keywords:** Effective Reproduction Number, Convex Optimization, COVID-19

## Abstract

COVID-19 is a global infectious disease that has affected millions of people. With new variants emerging with augmented transmission rates, slowing down of vaccine rollouts, and rising new cases threatening sanitary capabilities to the brink of collapse, there is the need to continue studying more effective forms to track its spread. This paper presents a strategy to compute the effective reproduction number *R*_*t*_. Our method starts with a form of the renewal equation of the birth process specially suitable to compute *R*_*t*_. After showing that one can express it as a linear system, we proceed to solve it, along with appropriate constraints, using convex optimization. We demonstrate the method’s effectiveness using synthetic and real sequences of infections and comparing it with a leading approach.

## Introduction

In December 2019, there was an atypical number of respiratory failure cases in Wuhan, China. By January 2020, laboratories could isolate the SARS-CoV-2, a new type of betacoronavirus^1^. Spread around the world, the SARS-CoV-2 has infected more than 100 million people, and more than two million people have died of COVID-19^2^. However, due to sub registration and asymptomatic patients, the actual numbers are likely to be higher. Research has established that the mean incubation time is five days; and that the serial interval, the time between symptoms onset for the infectious and infected, is between 4.1 and 7.0 days with a 95% confidence^3^. COVID-19 is a highly contagious disease, with a basic reproduction number, the average number of infections caused by an infected person when all population members are susceptible, for SARS-CoV-2 is around 2.24 and 3.58^4^.

Some researchers observe that a small number of cases (10 *∼* 20%) are responsible for a large number of infections (80 *∼* 90%)^5^, a significant fraction through asymptomatic or pre-symptomatic transmission^6^. However, super-spreaders act in super-spreader events, *i*.*e*., crowded and closed places where people may talk loudly, sing, shout to each other. These places include hospitals, leisure venues, workplaces, schools, family meetings, mass gatherings, prisons, homeless shelters, and transportation vehicles^7^. People with COVID-19 may develop symptoms that include cephalalgia, odynophagia, myalgia, arthralgia, rhinorrhea, dyspnoea, cyanosis, polypnea, anosmia, dysgeusia^8^. However, the definitive diagnosis comes after quantitative reverse transcription Polymerase Chain Reaction (RTq-PCR) tests^9^, which may take days to process. As new variants emerge with augmented transmission rates, vaccine rollouts slow down, and new cases threaten sanitary capabilities to the brink of collapse, there is the need to continue studying more effective forms to track infection spread.

During an epidemy, it is helpful to describe whether to apply lockdowns or permit certain activities. Early in the onset, when the whole population is susceptible, *R*_0_, the basic reproduction number, is a useful indicator of the epidemy’s virulence. Later in the process, it is of more practical value the effective reproduction number *R*_*t*_. *R*_*t*_ is the average number of infections that an infected person will cause. When *R*_*t*_ is one, conditions will remain the same. While with *R*_*t*_ *>* 1, the number of infected people will duplicate in a short time. On the contrary, if *R*_*t*_ *<* 1, the infection is receding and is tending to terminate.

This paper presents a strategy to compute *R*_*t*_. Our method starts with a form of the renewal equation of the birth process specially suitable to compute *R*_*t*_. After showing that one can express it as a linear system, we proceed to solve it, along with appropriate constraints, using convex optimization. We demonstrate the method’s effectiveness using synthetic sequences of infections and real sequences corresponding to cases in Mexico. We assess its performance using Cori *et al*.^10^, *a leading method of widespread use*.

### Related Literature

Due to its importance, researchers have explored various strategies to compute the effective reproduction number, *R*_*t*_ ^11^. One could classify these strategies between compartment-based, time series-based methods, and other methods.

### Compartment-based Models

Bettencourt and Ribeiro^12^ introduce a Bayesian scheme for the real-time estimation of the probability distribution of the effective reproduction number. Their approach develops a probabilistic formulation of the standard Suspected, Infected, Recovered (SIR) disease transmission model where the number of new cases is considered a stochastic discrete variable. At their end, Arenas *et al*.^13^ develop a method to estimate *R*_*t*_ based on an age-stratified compartment model. They require the transmission among individuals, specificities of a certain demographic group, and the human mobility patterns inside and among regions. Under assumptions of the infection probability, several contacts at each time step, and the fraction of susceptible individuals, they propose a Microscopic Markov Chain Approach (MMCA). Zhao *et al*.^14^ present a SIR vector-host compartmental model to compute *R*_*t*_ that is the product between *R*_0_ and the geometric average of the susceptibles of the host and vector populations. They found their approach consistent with past epidemics. Medina-Ortiz *et al*.^15^ use an autoregressive integrated moving average (ARIMA) model to forecast the number of infected *I*, recovered *R*, and dead individuals. Using these predictions, they estimate *R*_*t*_, based on a SIR model. Arroyo-Marioli *et al*.^16^ introduce a method to estimate *R*_*t*_ employing the growth rate of the number of infected individuals derived from the SIR model. However, the model seems to work well with the SEIR (SIR extended with a compartment for exposed) model. Their model combines epidemiology theory and time-series techniques in the form of the Kalman filter.

De Oliveira^17^ introduce a model where they constraint the model variables and parameters during the estimation process. Starting with a SIRDC (susceptible-infected-resolving-deceased-recovered) model, they construct a convex quadratic optimization formulation with linear and convex quadratic constraints. Their model permits them to trade-off accuracy versus smoothness of the estimates. Cao *et al*.^18^ compute *R*_*t*_ from the daily reported cases using a combination of the average latent period, average latent infectious period, and the logarithmic growth rate of the case counts. To predict the future epidemy profile, they employ a deterministic Susceptible-Exposed-Infectious-Recovered-Death-Cumulative (SEIRDC) model. Chong^19^ uses the same model as Cao *et al*. by dynamically estimating the value of *K*. Cislaghi’s method^20^ construct their formulation based on the observation that the number of cases increases by a factor of *R*_*t*_ every *d* days. Annunziato and Asikainen^11^ extend this method solving for *R*_*t*_ the difference of the exponential grow equation for two extremes in an interval. The Robert Koch Institute^21^ estimates *R*_*t*_ as the ratio between the number of infections for two non-adjacent intervals of four days.

### Time Series-based Models

Wallinga and Lipsitch^22^ show that different equations to estimate the observed epidemic growth rate and the reproductive number differ on their respective generation interval distribution. Furthermore, they demonstrate that using the mean of the generation intervals results in an upper bound to *R*_*t*_ for a given growth rate. Alvarez *et al*.^23^ introduce a variational model from the incident cases and the serial interval. Using a formulation of the renewal equation, linking the incidence with *R*_*t*_, they detect the initial exponential growth date. They then compute *R*_*t*_ by minimizing an energy function with considers the solution’s ability to reproduce the incident curve, the regularity of *R*_*t*_, and the adjustment of the initial value to an initial estimate of *R*_0_. Das^24^ propose a method that combines time series of recent cases at the population level with the time interval between infection of primary and secondary patients in family clusters. They feed the data to Walling and Lipsitch^22^. Kight and Mishra^25^ use the incidence time series and the generation time distribution. Although researchers often use the serial interval distribution as a proxy to the generation time distribution, in an illness such as COVID-19, the infections start before the onset of symptoms. They infer the generation time distribution from parametric distributions of the serial interval, a Normal function, and the incubation period, a Gamma function, distributions.

Cori *et al*.^10^ introduce a method that employs a Bayesian estimation for *R*_*t*_. Assuming a Gamma *prior distribution*, they employ a Poisson time of service model to generate the posterior, from where they employ Maximum Likelihood to compute *R*_*t*_. Lytras *et al*.^26^ extend Cori *et al*.^10^ adding imputations for missing data and adjustment for reporting delay. Menendez^27^ shows that Cori *et al*.^10^ method can be reproduced effectively using the ratio between the incidence at time t and the value at half the serial interval. *Na et al*.^28^ propose a probabilistic methodology where they predict the number of daily new infections from the daily death count. They base their quadratic programming approach on the probability distribution of delays associated with symptom onset and death reporting. They model this delay with a Gamma distribution. Bonifazi *et al*.^29^ relate *R*_*t*_ to the estimation of the doubling time, *i*.*e*., the time it takes to double the number of infected persons, assuming exponential growth. They obtain a closed-form solution and compute uncertainty by changing their assumption about the generation period length.

### Other Approaches

Rudiger *et al*.^30^ propose a method to estimate *R*_*t*_ based on the contact data extracted from mobile phones. They track de-identified cell phones’ position and declare a contact when two devices are within a short distance. In their method, nodes are devices, and edges are contacts. Interestingly, they found a better correlation between the contact index, a metric for number and heterogeneity of contacts, than their quantity. They represent heterogeneity using the moments of the number of contacts distribution. Lai *et al*.^31^ use phylogeny, the change of genomes in 52 samples of COVID-19, to estimate the evolutionary rate, times of the most recent common ancestor (tMRCA), and demographic growth. They inferred the effective reproduction number employing the birth-rate skyline model, a method that extracts the presence of rate shifts based on phylogenetic trees, and Markov chain Monte Carlo analysis.

### Computing *R*_*t*_

Assuming that individuals infected present a common generative interval distribution **w**^T^ = (*w*_1_, …, *w*_*T*_), *i*.*e*., the relative frequency of infections one day, two days, or *s* days before day *t* remains constant, one may express the expected number of infections 𝔼[*I*_*t*_] observed at time *t* as

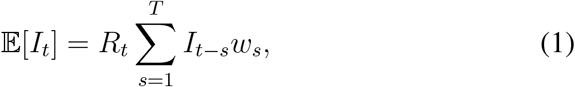

where one considers that the effective reproduction number *R*_*t*_ remains as if the conditions are as they were at time *t*, and *I*_*t*_ is the number of people infected at time *t*. The term **w** express a probability mass function and thus 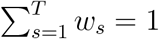 and *w*_*s*_ *≥* 0 for *s* = 1, …, *T*. Once we know *I*_*t*_, we may express (1) as

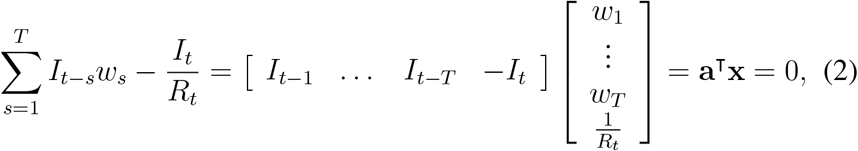

where **a**^T^ = [*I*_*t*−1_, …, *I*_*t*−*T*_, − *I*_*t*_] and **x**^T^ = [*w*_1_, …, *w*_*T*_, 1*/R*_*t*_]. To find the optimal **x**, one may express the dot product in (2) as a quadratic loss term such as

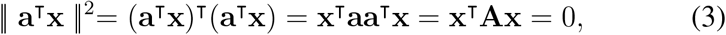

where **A**_(*T* +1)*×*(*T* +1)_ is a positive semi-definite symmetric matrix that has the form

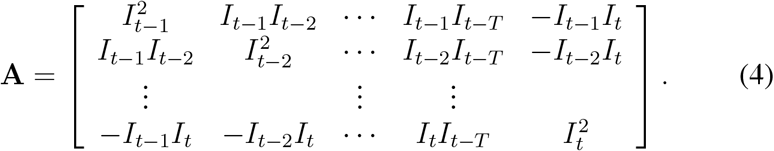

By construction, **A** is rank one, with its largest singular value equal to 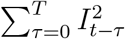; and nullspace with rank *T, i*.*e*., *T* of its singular values are equal to zero. We can now formulate the problem as a convex optimization using the standard formulation as

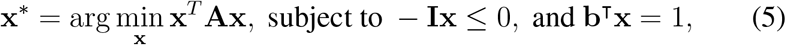

where **I**_(*T* +1)*×*(*T* +1)_ is the identity matrix, **b**^T^ = [1, 1, …, 1, 0]_1*×*(*T* +1)_ is a vector of *T* ones followed by one zero, *t ≥ T*, and *t*∈ ℕ, the set of natural numbers. Note that in (5) the objective function and the inequality constraints are convex, that is, if *f*_0_ = **x**^T^**Ax** and *f*_1_ = **Ix**, they satisfy

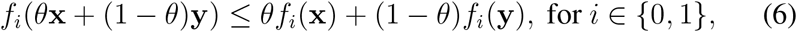

while the equality is affine; thus it has a global minimum^32^.

A possible strategy to solve (5) is through convex optimization. Fu *et al*.^33^ propose a novel strategy where first, they verify the cost function sign through disciplined convex programming (DCP). Then, they produce a standard form using graph implementation before finally applying a quadratic solver, such as OSQP^34^.

### Assessing Performance

To test our approach’s performance to compute *R*_*t*_, we relied on synthetic sequences made available by Gostic *et al*.^35^. They constructed their series based on an SEIR model, simulating an increasing level of contagion, followed by lockdowns to terminate in some release of activities (see Fig. 1)(a). We applied our model to this sequence using and compared with Cori *et al*.^10^. *In Fig. 1(b)*, we show in red the ground-truth value, in green the result of Cori *et al*.’s algorithm, and in blue the outcome of our method. We notice that after a slightly off start, with large uncertainty, Cori *et al*.’s method converges and faithfully follows the ground-truth value. Our approach follows the ground-truth value but, at inflection points, loses track and overshoots. However, it is worth observing that the optimization error remains low, with a largest value of 4 × 10^−11^, signaling that the method faithfully follows the infections curve. A by-product of our approach, the generative period distribution spreads without a significant mode or bias throughout the domain (see Fig. 1(c)), *i*.*e*., the contagious spreads equally over the generative interval.

**Figure 1.**
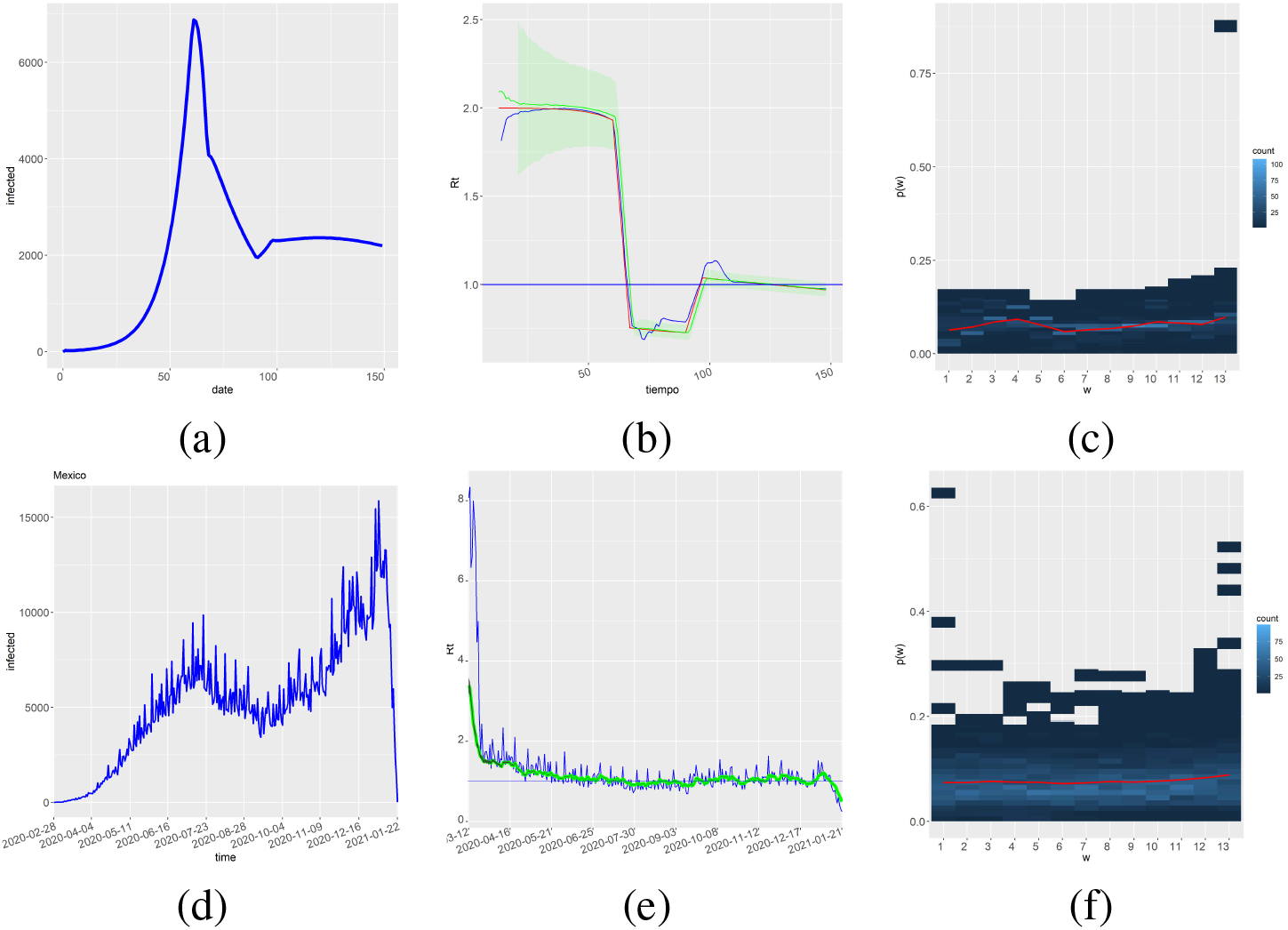
Assessment of our method in a real sequence. Using an SEIR model, Gostic *et al*. constructed a synthetic series of infections (a). We compare the result of our method (blue) with their ground truth (red) and the output of Cori *et al*. ^10^ (green) (b). Using the data provided by the Mexican Health Ministry, we processed the sequence of confirmed positives running from February 28, 2020, to January 22, 2021 (d). We compare the result of our method (blue) with the output of Cori *et al*. ^10^ (green) (e). Along with the result, we obtain the generative period distribution (c) and (f) for the synthetic and real sequence, respectively. The red line highlights the mean value position through the elements of **w**.

We then proceed to test our method using a real sequence of confirmed positives. To that effect, we took the COVID-19 dataset made public by the health ministry. In México, the authorities confirmed the first COVID-19 infection on February 28, 2020. Our sequence takes records until January 22, 2021 (see Fig. 1(d)). Before processing the information, we organize the sequence of infections by the onset of symptoms date and not by the date the authorities release the information. Thus, more recent dates will observe a reduced number of cases. We compare the output of our algorithm with Cori *et al*.^10^, in green in Fig. 1(e). We observe that our estimation tracks the spiking behavior observed in the input sequence, Cori *et al*. show a smooth development. However, as with the synthetic sequence, our method estimates an objective function error of at most 10^−10^ and a corresponding difference in estimating the next value *I*_*t*_ below | 2.5 × 10^−5^|. Finally, we observe in Fig. 1(f) that the generative interval’s data-driven estimation shows an even spread of the infection through time.

## Discussion and Conclusion

As a recent study justifies^35^, Cori *et al*.^10^ is regarded as a sound method to compute the effective reproduction number, *R*_*t*_. Cori *et al*.^10^ base their method in a Bayesian scheme that permits a variety of combinations between the priors and likelihood probability distributions. Our approach is data-driven and derives directly from a widespread definition for *R*_*t*_, requiring the determination of the generative period distribution length *T*. The optimization of our objective function results in a neglectable error, and since the resulting optimization function is convex, it has a single solution^32^. Other methods rely on assumptions on the nature of the priors and posteriors of *R*_*t*_, the transmission process dynamics. That may result in a more pleasant and smooth solution that deviates from the reported number of infections and possibly adding its own bias to the formulation. In this document, we introduce a novel method to compute the effective reproduction number, *R*_*t*_, based on the problem’s formulation as a convex optimization. Our experiments with synthetic and real data show that our method computes reliably *R*_*t*_ based only on the sequence of infected and the length of the generative period distribution.

Most infection reporting systems contain information delays affecting the number of cases input to the estimation of *R*_*t*_. In the future, we are planning to incorporate into our method a nowcasting estimation of the number of infections with its corresponding uncertainty assessment.

## Data Availability

To foster further research, allowing other researchers to verify our results and serve as a stepping stone, we make our code publicly available at \url{https://github.com/joaquinsalas/computingRt}.

https://github.com/joaquinsalas/computingRt

## Acknowledgements

Thanks to the Mexican Health Ministry for the data provided and to the CDMX SECTEI discussion group.

## Author biography

Joaquín Salas is a professor in the field of Computer Vision at Instituto Politécnico Nacional. Member of the Mexican National Research System, his research interests include the monitoring of natural systems using visual perception and aerial platforms. Salas received a Ph.D. in computer science from ITESM, México. He has been a visiting scholar at Stanford University, Duke University, Oregon State University, Xerox PARC, the Computer Vision Center, and the École Nationale Supérieure des Télécommunications de Bretagne. He has served as co-chairperson of the Mexican Conference for Patter Recognition three times. Salas was Fulbright scholar for the US State Department. He has been invited editor for Elsevier Pattern Recognition and Pattern Recognition Letters. For his services at the Instituto Politécnico Nacional, he received the *La’zaro Ca’rdenas* medal from the President of Mexico.

## Declaration of conflicting interests

The author declared no potential conflicts of interest with respect to the research, authorship, and/or publication of this article.

## Funding

This work was partially funded by SIP-IPN 20210219.

## Supplemental material

To foster further research, allowing other researchers to verify our results and serve as a stepping stone, we make our code publicly available at https://github.com/joaquinsalas/computingRt.

## Notes

### Competing Interest Statement

The authors have declared no competing interest.

### Author Declarations

no needed for this work

